# A Systematic Review of Military to Civilian Transition: The Role of Gender

**DOI:** 10.1101/2024.02.22.24303195

**Authors:** Alexandria Smith, Laura Rafferty, Bethany Croak, Neil Greenberg, Rafiyah Khan, Victoria Langston, Marie-Louise Sharp, Anne Stagg, Nicola Fear, Sharon Stevelink

## Abstract

**Background:** The military-to-civilian transition can be a challenging period for many service members; however, recent research suggests that female ex-service personnel (veterans) confront additional complexities during reintegration into civilian life. This systematic review aimed to identify and synthesise findings across qualitative studies exploring the impact of gender on this transition process.

**Methods:** Peer-reviewed literature was drawn from a multi-database search, limited to qualitative studies. The studies included either female veterans or both male and female veterans aged 18 years or older who had previously served in the Armed Forces within the Five Eyes (FVEY) countries (Australia, Canada, New Zealand, the United Kingdom, and the United States). We used a Framework Analysis approach to guide the synthesis of the qualitative data. An assessment of study quality was conducted using the Joanna Briggs Institute (JBI) Qualitative Critical Appraisal Checklist for Qualitative Studies. The study protocol is registered with the Open Science Framework (osf.io/5stuj).

**Results:** In total, 10,113 articles were screened after the removal of duplicates, 161 underwent full-text review, with 19 meeting the eligibility criteria. The review identified eleven themes split across individual’s experience whilst serving and after transitioning out of the military service. Both male and female veterans discussed a period of acculturation when they joined service and adapted to military norms, culture and identity. Female veterans faced additional challenges at this stage centred on the conflict between feminine norms and the military masculine ideal. Upon leaving service both male and female veterans experienced a loss of military identity and purpose, and dissonance with civilian norms illustrating a military-civilian divide. For female veterans, adjustments and adaptations learned in the military clashed with civilian feminine norms and stereotypically male veteran culture. Female veterans also struggled with the legacies of gender inequality, discrimination, and sexual assault which affected their development of a female veteran identity and affected the provision of services designed to meet their needs as a female. Despite these challenges, female veterans’ expressed pride in their service and accomplishments.

**Conclusions:** Any effort to improve the military-to-civilian transition should take account of the legacy of gender discrimination, especially within the military service, and the potential mismatch between historical civilian female norms and the more traditionally masculine norms of military life.

**Disclosures:** This project was supported by a grant from the Forces in Mind Trust (FiMT) 2202. Full ethical clearance was granted by the Health Faculties Research Ethics Subcommittee, King’s College London. Project Reference: HR/DP-22/23-33303.

## Background

Transitioning from the Armed Forces (AF) to civilian life can be challenging for many ex-service personnel (veterans), with many veterans reporting some difficulty integrating into the civilian environment ^1^. The loss of their military identity, and the attendant isolation following separation from their military community, can impede service members from fully integrating, or accessing support ^2–4^. A negative transition out of the military is associated with a number of undesirable outcomes including economic and housing instability, family conflict, poor physical and mental health, and reduced well-being ^5^. In contrast, the adaptation of one’s military identity to civilian cultural values and norms has been shown to improve the transition experience ^6–9^.

The number of women in the AF is projected to increase over the next decade with the expansion of roles available to women and targeted recruitment and retention efforts ^10–14^. Currently, women account for 11% to 18% of the regular AF within the Five Eyes (FVEY) (Australia, Canada, New Zealand (NZ), the United Kingdom (UK), and the United States (US)) ^10,11,13–15^. Despite the increase in women serving, most of the research exploring the transition process has focused on men ^16^. Yet even with the lag in research among female compared to male veterans, this is quickly becoming a burgeoning research area ^17^. Much of the current evidence is drawn from the US and suggests female veterans confront additional complexities in the military to civilian transition compared to male veterans and experience worse physical and mental health outcomes ^17–19^. The experience from other FVEY countries is less clear ^20^. Governments in the FVEY countries recognize the importance of improving the transition experience for male and female veterans and have begun implementing several policies to support transition ^10,21–24^. However, there remains ample room for improvement in policies and programs to support female veterans and in order to do this effectively we need further understand how women’s military experiences and post transition experiences may differ compared to those of men.

A previous scoping review of the role of gender in the military to civilian transition primarily identified quantitative studies related to physical and mental health outcomes ^25^. The authors of this scoping review noted a rapid rise in the number of publications across their period of inclusion (1990-2015) ^25^. In contrast, the proposed study will conduct a synthesis of the current *qualitative* literature examining the *experience* of military to civilian transition, and the impact of gender on this transition.

## Methods

### Information Sources and Search Strategy

A comprehensive search strategy was designed in consultation with a data librarian. The study protocol was registered with the Open Science Framework (osf.io/5stuj) ^26^. A multi-database search was conducted, including Medline, Embase, PsycINFO, Pubmed, Global Health, Web of Science, and EBSCO. The search strategy included controlled vocabulary terms and keyword searches using variations of the following search terms; ‘females,’ ‘veterans,’ and ‘transition’. Additional detail regarding the search terms and the number of articles retrieved is provided in Appendix 1. Reference sections of articles were also cross-checked for additional articles of relevance. Our original comprehensive search was conducted in February 2023 and an update search in February 2024 to ensure that the results reflected the most current evidence.

### Inclusion and exclusion criteria

Papers were included if they were peer-reviewed, qualitative or mixed methods studies, and included female participants, age 18 years or older, who had previously served in the AF within the FVEY (Australia, Canada, NZ, the UK, and the US). The definition of ‘veteran status’ was determined by the individual authors and these definitions likely varied by across nations ^27^. Papers were excluded if they reported solely on male participants or where gender was not identifiable, were not written in English, or were inaccessible in full text at the time of the review. There was no restriction placed on the publication date.

### Study Selection

Duplicate studies were removed first by EndNote ^28^, followed by manual checks. The selection of studies occurred in three stages: title; abstract; and full-text review. Initial screenings of titles and then abstracts were conducted by one reviewer (AS). Two reviewers (AS, LR) independently conducted full-text reviews with consensus required for study inclusion.

An initial search was run in February 2023, and an updated search was conducted in February 2024, which resulted in 10,113 articles after the removal of duplicates. The title and abstract review excluded 9,952, leaving 161 for full text review. Of these, 19 studies met eligibility criteria (Figure 1. Prisma 2020 Flow Diagram). Two reviewers (AS and BC) independently conducted an assessment of study quality using the Joanna Briggs Institute (JBI) Qualitative Critical Appraisal Checklist for Qualitative Studies ^29^ which is provided in Appendix A.

**Figure 1.** Prisma 2020 Flow Diagram

### Data Extraction

Key study details were extracted into a standardized form, including author(s), year of publication, country, participant characteristics, interview type and analysis. A summary of extracted information is provided in Table 1.

**Table 1:** Primary characteristics of studies included in the systematic review.

### Synthesis

A Framework Analysis approach was used to guide the synthesis of the qualitative data extracted from the studies ^30^. Relevant data was drawn from the original authors’ interpretation of the results, the discussion section, and direct citations from participants.

Framework Analysis involves the development of a matrix output where rows and columns are used to summarise cases (in this case qualitative studies) and codes to summarise data. A Framework Analysis typically follows seven stages, the latter six of which were utilized in this research: transcription of verbal data to text; familiarisation; coding; developing an analytical framework; applying the analytical framework; charting data into the matrix; interpreting the data ^30^.

Each study included in the review was read for familiarisation with comprehensive notes taken. The *Results* and *Discussion* sections were carefully read line by line with codes (para-phases or labels) assigned to text as appropriate. During this process, an iterative approach was employed with frequent discussions among researchers to ensure consensus in the interpretation of the findings. After coding several papers, LR and AS met and compared codes to create a unified set of codes and to develop a tree diagram to group the codes together. Several iterations of this analytical framework were created until all the studies included in the review had been analysed. Once the framework was finalized, studies were re-read, indexed, and charted onto a matrix displaying the analytical framework. The results of the analysis were explored, and broader concepts around the process of transition were developed.

## Results

Nineteen papers were included in the full review with the characteristics of each study detailed in Table 1. No studies were excluded due to concerns with quality (Appendix A). Most studies were from the US (n=14) ^2,3,7,31–41^; one study included participants from both the US and Israel ^42^. The UK contributed two studies ^43,44^ with Canada and Australia each with an additional study ^45,46^. A total of n=502 veterans were included in the qualitative studies, of which n=290 were female. The participants ranged from 22 to 71 years of age, with a length of service between 2 to 30 years. The military characteristics of studies varied, including rank (enlisted and officer), branch, and occupational specialties. Most female participants had been deployed at least once. Studies were published from 2013 through 2024, with more than half of the studies (n=11) published after 2020.

The included studies implemented a variety of analytical frameworks which informed their interpretation of the qualitative data. The most frequent analytic frameworks used included thematic analysis (n=9) ^2,3,31,33,35,40,46–48^, grounded theory (n =5) ^36–39,45^, and phenomenological methodology (n=6) ^7,32,34,41,44,47^. Studies also employed a critical feminist approach (n=1) ^49^ or feminist narrative analysis (n=1) ^42^. The studies generated n=80 themes and n=91 subthemes exploring the transition process between the military and civilian environment and the role of gender during military service and post-service. Language regarding gender and its role in transition is largely taken from the studies themselves noting that gender incorporates cultural, social, and institutional meaning prescribed to one’s sex. A discussion of feminist theory is beyond the scope of this review; however, we acknowledge the vast and evolving theories of gender, particularly how gender is operationalized within a military institution ^49,50^. The interrelation of themes and subthemes is provided in Table 2. Explicit detail of the themes and subthemes extracted is available in the appendix (Appendix B).

**Table 2:** Qualitative themes identified in the systematic review.

## Study findings

### In-service Military Experience

#### Indoctrination: Military identity and norms

Following enlistment in the military, service members undergo an intensive process of indoctrination ^45,46,51^. Participants describe an initial “culture shock” as they adjusted to the new military envionnment^37,38^. Military norms are introduced through the emotionally and physically intensive training, where all individuals are expected to cast off effeminate qualities and exhibit the desired, stereotypically, masculine norms ^32,52,53^. Studies describe how both men and women must adjust to the hegemonic masculine norms of the military^45^. Prized qualities reported included aggressiveness, physical and emotional toughness, dominance, bravery, competence, service to fellow soldiers, loyalty, and technical expertise^51,53,54^. Womanhood, femininity, and non-heteronormativity were reported to be seen as weak and subordinate to military hegemonic masculine norms ^31,54^. Service members shifted their values and beliefs to reflect those supported by the military institutions, which were in turn, reinforced and solidified through their tenure in the military and the execution of their military duties^51^.

#### Impact of gender

##### Feminine at odds with masculine ideal

Servicewomen in the studies described their gender as being in direct conflict with the stereotypically masculine norms of the military. They were forced to reconcile their identity as a female, a soldier, and for some a partner and mother, which were potentially in tension with one another ^52^. Women in the studies described a variety of strategies used to adapt to a highly masculine military environment, including taking on the more masculine identities of their male counterparts, reducing outward feminine mannerisms, changing the way they moved, adjusting the tonality or content of speech, and being permissive of, or even joining in with the dominant humour ^7,32,38,44,55^. Despite these efforts, many women expressed feeling unable to fully integrate and noted the stress of constantly adjusting themselves to fit the masculine norm ^4,27,42,45^. Eichler (2022) noted that as women attempted to assimilate by reducing their femininity, they were paradoxically “hyper-visible” as a minority member in the Armed Forces^45^, highlighting the “otherness” of being female which remained a barrier to attaining full group membership ^45^.

##### Gender inequality, misogyny and discrimination

Gender inequality, gender-based discrimination, and hostile work environments where men are viewed and treated as superior, were described as pervasive within the Armed Forces ^7,31,32,38,45^. Many women voiced the need to work harder than their male counterparts to be perceived as valued members of the military ^7,31,32,42,45^. Women experienced continuous pressure for high-level performance in a bid to be seen as equal to men ^7,45^. Mistakes were seen as confirmation of their inadequacy and reflected poorly on other female service members ^7,45^. Some women reported that they were assigned menial tasks and their career progression was constrained because of their gender ^45^. Gender inequality was also apparent in the lack of physical preparedness in the military for female service members with military equipment and clothing provided to women deemed inadequate to meet their needs ^31,45^. These items were often ill-fitting and unsuited to the female statue, often resulting in injury ^31^. These biases further extended to gender-specific health needs with inadequate health services and medical expertise to manage genitourinary conditions and reproductive health concerns specific to women ^12,31^.

Female service members also experienced sexual objectification, sexual harassment and the threat of sexual assault ^31,39,42,45^. These threats increased during periods of deployment, requiring a heightened sense of awareness and compounding trauma experienced by service members whilst on deployment ^31,42^. While both discrimination, harassment and assault were reported as pervasive across the studies, women noted differences depending on a history of deployment, service branch, rank, employment, and ethnicity ^38^. For example, female service members working in the healthcare sector and women of higher rank reported discrimination to a far lesser degree than women of lower rank, women in forward-facing or combat positions, and women of colour ^31,32,38^.

### Post-Service Experiences

#### Loss of military identity

For many service members, whether male or female, transitioning out of the military resulted in a profound sense of identity loss and required a renegotiation of one’s identity within civilian society ^35,41,44,47^. Re-entry to civilian life was often accompanied by a “reverse culture shock” as ex-service members constructed a new post-military identity and adjusted to the norms and expectations of civilian life ^37,47^.

A portion of male and female veterans expressed a sense of bereavement at losing their military identity and struggled with constructing a new identity in its place ^2,7,32,41,47^. Some veterans described living two separate lives -- one military and one civilian ^7,38,47^. At one end, veterans actively fostered their military identity and rejected any alignment with a civilian identity ^34^. On the other, veterans leaned into the civilian identity, over time fading their military identity ^34^. However, most male and female veterans in the included studies occupied a middle realm, balancing their military identity with a civilian identity while constructing a new veteran identity ^7,32,34,38,40,41^.

#### Military – Civilian divide

Both male and female veterans routinely described the large chasm between military and civilian norms^2,7,35,37,40,44^. Veterans described feeling unable to be themselves around civilians due to differences in dress, mannerisms, humour, language and being perceived as inappropriate or aggressive ^2,33,47^. These differences in military and civilian social norms impeded upon building meaningful relationships with civilians ^2,7,33,35,37,46,47^. Some veterans found the detachment from civilians so profound that they were unable to reconnect with previously meaningful networks, such as childhood friends ^38,47^. The lack of social connection left veterans detached and isolated from both their military and civilian community, often leaving them to navigate the hurdles of transition alone ^2,7,35,37,46,47^.

In workplace settings, the tension between military and civilian norms was especially stark. Both male and female veterans felt their unique skills were underutilized and underappreciated in the civilian workforce ^2,33–35,40,56^. The unstructured nature of work, lack of clear policies, operating procedures, or clean lines of responsibility frustrated some veterans ^36,38^. Conversely, civilian employers often perceived veterans as rigid and inflexible in their working style. Veterans were also frustrated by their work colleagues and felt many lacked accountability, discipline, punctuality, and motivation ^38^. Many veterans found civilian employment mundane, slow-paced, and the civilian workforce lacking motivation ^3,35,37,40,42,56^. These sentiments were even evident in fields with equivalents in the civilian sector, such as healthcare ^2,37^.

#### Loss of purpose

Many male and female veterans described a general lack of purpose or meaning in the civilian environment ^2,33,35,37,40,41,44,46,56^. Veterans described the mismatch between the slower pace of civilian life set against the fast pace of military life ^37^. Some veterans objected to the lack of urgency displayed by civilians, even family members, and were frustrated by the absence of resolve, structure and routine in their day-to-day lives^2,35^. Paradoxically, veterans also commented on feeling overwhelmed by the amount of choice, fluidity, and lack of central hierarchy in the civilian environment ^2,35,38^. Many veterans found executing general activities of daily living, such as appointments or errands, to be overwhelming ^35^.

### Impact of gender

The way women left the military impacted their post-service identity formation. Some female veterans felt forced out due to a lack of upward mobility, being side-lined after family leave, or when demands as a parent conflicted with the needs of the military ^45^. Other women left following sexual discrimination, harassment, or assault. Those who left the service earlier than desired often expressed feelings of betrayal from the military ^7,32,35,38,42,45^.

#### Feminine norms

Gender norms were described as imposing an unnecessary burden on women with many female veterans experiencing a discordance between feminine civilian and military norms ^7,38,45^. The adjustments women made to adapt to the male-dominated military environment were often less well received in civilian roles^38^. The expression of stereotypical feminine norms in the civilian environment, communicated through clothing, physical movement, mannerisms, and tonality of speech, informed all levels of interactions^38,45,47^. These included social rules for relationships, family dynamics, and workplace environment^45,47^. Female veterans described being perceived as aggressive or domineering^38^. These tensions were further complicated for women who had deployed or engaged in combat roles, as these experiences were at odds with the feminine ideal in civilian society^38,45^.

Feminine norms associated with divisions of labour continued to apply pressure on women serving in the military, with the role of caregiving disproportionately falling to women. Just over half of all partnered female service members were in a dual-serving household, with family demands often cited as a predominate catalyst for exiting the armed forces ^45^. Given the elevated pressures and demands, partnerships often ended in separation ^45^.

#### Female veteran identity and needs

Female ex-service members were not able to as easily embrace a veteran identity compared to their male counterparts ^45^. Women noted the inherent tension between their multiple identities as warrior, spouse and mother compared to the more concordant masculine identity of warrior, provider, and father ^45^. These sentiments were also confirmed through their interactions with civilians and even healthcare providers. Female veterans noted a lack of recognition of their military service or a minimization of their role in the military by both civilians and military healthcare providers ^31,39,45^.

Women veterans noted that healthcare and support services specific to their needs were routinely unaddressed ^31,39^. They were often dismissed when seeking care for misunderstood chronic conditions, including fibromyalgia, chronic fatigue syndrome, and genitourinary concerns ^31,39^. The potential connection between these conditions and military service were often underrecognized or negated ^31^. Women also reported several reproductive challenges that they related to their exposures during deployment ^31^. Women noted that these emerging and chronic conditions, such as urogynaecological, fatigue, migraines, and neurological disorders were often dismissed or misunderstood even among providers familiar with the veteran population (e.g. Veterans Affairs, US) ^31,39^.

#### Consequence of Military Sexual Trauma

The negative consequences of Military Sexual Trauma (MST) were prominent themes that emerged across studies ^7,31,39^. While no study directly asked about past experiences of harassment or assault, many female veterans voluntarily shared their experiences. The prolonged negative effect of MST extended beyond veterans mental and physical health but also affected their personal, social, and professional spheres ^2,7,31,36,38,39,42,56^. Female veterans spoke about the adverse mental health effects, the avoidant behaviour, irritability, depression, anxiety, and PTSD resulting from MST ^7,31,39^. Women also spoke of the lack of accountability, retaliation from reporting, and the sentiment that the military placed unit cohesion above the safety of female soldiers, often pushing women out of military service ^36,38,34^. Of note, no male veteran participants provided accounts of sexual violence. Sexual violence and assault occur at a higher prevalence among female service members compared to male service members, however, male service members are less likely to report sexual violence ^57^.

## Discussion

### Overview

The review of the qualitative literature examined the transition experience among male and female veterans to understand the potential effect of gender on this transition. While many veterans who transition from military service to the civilian environment report modest to minimal difficulty; a substantial portion of veterans face hardship during both the short and long-term ^1^. A loss of military identity, a reduced sense of meaning and purpose, and social isolation relative to the military-civilian divide were prominent and resounding themes for all veterans, with female veterans reporting that that their gender amplified these challenges as discussed below.

### Identity instability

For male service members, adopting a military identity entailed assuming a stereotypically hyper-masculine form. When they then transitioned out of the AF, this hyper-masculinity was still accepted and congruent with male civilian norms and expectations of male ex-service members. On the other hand, the formation of a military identity for female service members occurred in an environment which often contested their presence. To mitigate this, female service members would often adopt a more stereotypically masculine persona. However, as female service members transitioned to the civilian environment, they often found that their military identity and the adjustments they had made to fit the military environment were now in misalignment with societal norms and expectations for civilian women. Thus, women found themselves in a perpetual process of identity formation, deconstruction, and renegotiation, in response to institutional and social norms and expectations whilst adapting from civilian to military and then again back to civilian environments.

The constant negation of identity by the external environment and renegotiation of one’s identity may have detrimental effects on female veterans’ physical and mental health and overall wellbeing ^58–60^. For female veterans, their identity as a military service member and subsequently a veteran was often contested by their primary social environments, either in overt or subtle ways. While the theoretical underpinnings of individual and social identity are complex, nuanced and beyond the scope of this paper, studies from disparate disciplines all emphasise the importance of a coherent identity in supporting mental and physical wellbeing, creating meaning and purpose, combating loneliness, and fostering social cohesion ^61–64^.

### Gender discrimination

Women’s transition experiences were also affected by gender-based discrimination, harassment and assault, and its enduring effects. Gender-based discrimination and harassment is undeniably corrosive in the workplace^65^. Negative consequences include the erosion of career advancement, reduction in job satisfaction, retention, and motivation, and worse overall performance ^66,67^. These negative consequences are reflected in the qualitative studies assessed. Women articulated the challenges they faced in performing their duties due to the presence of discrimination and harassment. Women in the studies also reported an increase in gender-based discrimination, harassment, and violence during deployments. Many independently described the added vigilance required to maintain their safety both from enemy combatants as well as against fellow service members.

Women in the qualitative studies described feeling pushed out of the military, leaving out of necessity rather than choice. Some of these women expressed feeling betrayed by the military institution and noted the role of institutional barriers which resulted in the need to exit. Taking into consideration accumulated stressors, including the demands of caregiving, experiencing discrimination or harassment, physical injuries, or being a victim of assault, it is not unexpected that women often have shorter tenures in the military compared to their male counterparts and are more likely to leave before completion of their terms of service ^24,68^. Of concern, early service leavers (ESLs), tend to have worse mental health outcomes compared to individuals who complete their service contract ^69,70^. Studies of workplace harassment have demonstrated that negative institutional responses contribute to long term adverse psychological harm ^71^.

### Positive sentiments

Yet, despite challenges noted in the review, many women felt a sense of accomplishment being able to perform within the male-dominated environment ^38,42^. Females service members took pride in their ability to complete exacting physical standards and saw their military identity as hard won ^32,38,42^. Women often formed strong bonds within the military, despite experiencing various forms of exclusion ^7,31,47^. Some women referred to their fellow service members as family and cited loyalty, duty, and respect for their fellow soldiers as reasons for social cohesion ^36,47^. These accomplishments often buoyed women in the transition period, allowing them to feel more capable in managing civilian demands ^39^. These positive emotions matter.

### Major gaps

There is an emerging body of evidence examining the military to civilian transition among women ^17^. However, numerous gaps in understanding remain. From the reviewed studies, there are three areas where knowledge is notably lacking: the role of a military and veteran identity among women, the long-term detrimental effects of discrimination and harassment, and the evaluation of health needs and services specific to female service members and female veterans.

For men and women, a sense of identity supports self-esteem, provides meaning and purpose, and connection with a wider social group. As such, changes in identity or conflicts between one’s identity and the surrounding social norms and values can manifest in several negative ways, such as reduced mental or physical wellbeing. The development and role of personal identity for women during their military service and transition needs to be further understood. Second, given the pervasiveness of gender-based discrimination and harassment, the long-term social, mental, physical, and economic costs to service members should be examined. It is critical to understand how discrimination based on any characteristic, such as gender, ethnicity, or religious affiliation, may undermine force readiness and unit cohesion. Third, a more comprehensive assessment of the diverse social and health needs and enhanced services is required. Female service members and veterans spoke to a wide range of physical and mental health needs that vary according to individual needs and preferences, life stage, and environment. A broader conceptualization of veteran women’s health will be required to address these needs effectively. This understanding should encompass concerns such as reproductive health, perimenopause and menopause, cardiovascular disease, cancer, autoimmune disorders, and mental health ^72,73^.

### Strengths and Limitations

This review’s strengths include a comprehensive search strategy developed in collaboration with a data librarian. An expanded set of controlled vocabulary terms and keywords were used across several databases to ensure relevant studies were identified. The review builds upon previous quantitative work where the synthesis of qualitative data allows for a richer understanding of the female veteran experience by finding the commonalities and differences in the veteran experience. There are some limitations with the findings of the review. First, most of the studies were from the US thus potentially limiting the generalizability of the female veteran experience to other countries. Second, there is a diversity of approaches to qualitative synthesis each with inherent limitations and biases. We included all studies, regardless of differing underlying methodology (phenomenology, grounded theory, narrative analysis, etc.). Third, the preponderance of studies demonstrated significant difficulties during and post service, with gender prominently influencing these difficulties. While there was considerable similarity in themes across the studies, these experiences may not encompass all of the experiences of female veterans.

## Conclusion

This review found that female veterans confront additional stressors when leaving the Armed Forces compared to their male counterparts. The majority of studies found female service members reported experiences of varying degrees of gender-based discrimination, misogyny, and gender inequality both during and post-service with MST continuing to be a concern. As women continue to enter the AF in greater numbers over the coming decades, research must advance our understanding of the barriers to a successful transition for female service personnel, particularly beyond the US context. For advancement, policymakers and the Armed Forces must also be willing to confront the cultural legacy and entrenchment of gender inequality, discrimination and MST.

## Authors contributions

1. Conceptualization: LR, BC, NG, SS, NF
2. Data curation: AS
3. Formal analysis: AS, LR, RK, BC
4. Funding acquisition: LR, BC, NG, SS, NF
5. Investigation: AS, LR, RK, BC
6. Methodology: AS, LR
7. Project administration: AS
8. Resources: NF
9. Software: AS
10. Supervision LR, MLS, SS, NF
11. Validation: AS, LR, BC, RK
12. Visualisation: AS, LR
13. Writing (original draft) AS, LR, BC
14. Writing (review & editing) AS, LR, BC, NG, RK, VL, MLS, AS, NF, SS

## Ethics

This project received full ethical clearance granted by the Health Faculties Research Ethics Subcommittee, King’s College London. Project Reference: HR/DP-22/23-33303

## Data Availability

All data produced in the present study are available upon reasonable request to the author.
Smith, A. (2023, November 30). A Systematic Review of Military to Civilian Transition: the Role of Gender. Retrieved from osf.io/5stuj

