## Supplementary material for "A Systematic Review of Military to Civilian Transition: The Role of Gender": Table 1 (from another version of this preprint: version 2, 2024-04-17)

| Table 1: Primary characteristics of studies included in the systematic review | | | | | |
| --- | --- | --- | --- | --- | --- |
| Author Publication Year | Country | Participant Characteristics | Primary Underlying Theories | Method of Inquiry | Type of Qualitative Analysis |
| Raabe et al. (2024) | USA | Women (n=3), Men (n=13)  *Characteristics for women only*  Age Range: 31-34  Length of service: 8-12  Months since discharge: 3-28  Branch: Army (n=2), Marines (n=1), | Self-determination theory^1^ | Semi-structured in-depth interviews | Thematic analysis |
| Barrington et al. (2023) | UK | Women (n=1), Men (n=4)  *Characteristics of full sample (men and women)*  Age Range: 35-55  Branch: Army (n=5)  Length of service: 5-20+ years  Years since discharge: <1 – 20 | Social identity theory^2^ | Semi-structured in-depth interviews | Phenomenological methodology |
| Murray & Cancio (2023) | USA | Women (n=1), Men (n=5)  *Characteristics of full sample (men and women)*  Age Range: 30-55 years, average = 42.7  Branch: Army (n=4), Navy (n=1) Marine (n=1)  Length of service: 5- 28 years, average 12.6 years | Community reintegration^3^ | Semi-structured in-depth interviews | Phenomenological methodology |
| Rattray et al. (2023) | USA | Women (n=8)  Age Range: 24-53 years, average = 34  Years since discharge (average): 34 | Discovery-oriented approach^4^ | Semi-structured interviews at baseline, month 6,12,18 and 24 | Grounded theory, inductive thematic analysis |
| Barnett et al. (2022) | Australia | Women (n=6) Men (n=34)  *Characteristics of full sample (men and women)*  Age Range: 25–57, average = 37  Length of service: 4-40 years, average = 15.1  Branch: Army (n=25), Navy (n=7), Air Force (n=6), Army and Airforce (n=1), Army and Navy (n=1)  Reason for leaving discharge: Own request (n=24), Medical discharge (n=11), Compulsory age retirement (n=1), Other (n=4) | Social identity theory ^2^ and Self-categorization theory ^5^ | Semi-structured in-depth interviews | Thematic analysis |
| Eichler (2022) | Canada | Women (n= 33)  Age Range: 27–64, average = 47 Length of service: 2 – 39.5 years, average = 19 years Branch: Air Force (n = 10), Army (n = 7), Navy (n = 5), combination (n = 11) | Critical feminist approach with gender as primary category of analysis ^6,7^ | Semi-structured, in-depth interviews and focus groups | Grounded theory and critical feminist approach |
| Guthrie-Gowerm & Wilson-Menzfeld (2022) | UK | Women (n=5), Men (n=6)  *Characteristics for women only* Age range 41-72, average 55.4 Length of service: 7-30 years, average 14.4 years Years since discharge: 15-27 years, average 18.8 years | Loneliness through the Social Needs Approach  and the Cognitive Discrepancy Model^8^ | Semi-structured in-depth interviews | Phenomenological methodology and thematic analysis |
| Laferty et al. (2022) | USA | Women (n =10) Age Range: 51-85, Average = 59 Branch: Army (n=7), Army National Guard (n=1), Marine Corps (n=1), Air Force (n=1) | None stated | Semi-structured in-depth interviews | Content analysis and inductive thematic analysis |
| Boros et al. (2021) | USA | Women (n=4) Age Range: 40-60, average = 46.5 Branch: Army (n=2), Air Force (n=2) | Goffman’s Total institution ^9^; Overarching Feminist Theory^10^ | Semi-structured, in-depth interviews | Interpretative phenomenological analysis |
| Daphna-Tekoah et al. (2021) | USA / Israel | Women (n=20), United States military (n=10), Israel Defence Forces (n=10) All women completed combat or combat support service | Interdisciplinary: Feminist Security Studies (FSS), International Relations, Guide to Listening^11,12^ | Semi-structured, in-depth interviews ^†^ | Feminist narrative analysis |
| Sayer et al. (2021) | USA | Women (n=50), Men (n=50).  *Characteristics of full sample (men and women)* Average age = 40.1  Military component: Active duty (n=50), Reserve/Guard (n=50) Military Branch: Army (n=58), Air Force (n=19), Navy (n=16), Marine Corps (n=7) Rank*:* Enlisted (n=83), Officer (n=17) | Belonging and social connectedness ^13^; Ecological model^14^ - interaction between individuals and social environment | Written narrative | Thematic analysis of written essays. |
| Leigh & Koblinsky (2017) | USA | Women (n= 29)  Branch: Army (n=19), Navy (n=5), Air Force (n=3), Marine Corps (n=2)  Rank: Enlisted (n=20), Officer (n=8) Deployed (n=22) | Organismic Valuing Theory of Growth through Adversity^15^ | Focus groups with semi structured interviews | Thematic analysis of focus groups |
| Libin et al. (2017) | USA | Women (n=2), men (n=8)  *Characteristics for women only* Age: 28 and 32 Branch: Army (n=1), Air Force (n=1) | Schlossberg's Transition Theory ^16^ ; Role exit ^17^ | Semi-structured, in-depth interviews | Phenomenographic approach |
| Orazem et al. (2017) | USA | Women (n = 42), Men (n = 58) *Characteristics of full sample (men and women)*  Age: range 22 - 66 years, avg. 36  Branch: Army (n=60), Air Force (n=16), Navy (n=13), and Marine Corps (n=11) Rank: Enlisted (n=82), officers/warrant officers (n=18) | Theories of identity formation ^18^  Erikson’s stages of psychosocial development ^19^ | Written narrative describing reintegration difficulties | Thematic analysis |
| Ahern et al. (2015) | USA | Women (n=7), Men (n=17)  *Characteristics of full sample (men and women)* Age range: 22 - 55.  Branch: Air Force (n=2), Army (n=8), Army National Guard/Reserves (n=5), Marines (n=5), Navy (n=4) Time since discharge: 75% within the last 4 years | Homecoming Theory^20^ | Semi-structured, in-depth interviews | Thematic analysis |
| Burkhart & Hogan (2015) | USA | Women (n = 20)  Age range 23-65 years, average 45 years Branch: Navy (n = 7), Air Force (n = 6), Army (n = 5), Marines (n = 2). Rank: enlisted (n=10), Commissioned officers (n=10)  Length of Service 2–30 years, average 16.7 years. | Erikson’s stages of psychosocial development^19^ | Semi-structured, in-depth interviews | Grounded Theory |
| Mankowski et al. (2015) | USA | Women (n=18) Age range: 27-63, median age 43.5  Rank: E 3-5 (n=6), E6-9 (n= 8), O 3 - 5 (n= 4) All had been deployed 2+ times. | Bridge theory: military as a bridge to adult roles ^21,22^ | Semi-structured, in-depth interviews | Grounded theory, constant comparative method |
| Koenig et al. (2014) | USA | Women (n=14), Men (n =17)  *Characteristics of full sample (men and women)* Age: Median 30 Rank: Enlisted 20 (64.5%), Officer 11 (35.5) All had been deployed to Iraq or Afghanistan. | Faulkner: Stages in the re-entry from military to civilian life^23^; | In person semi structured interviews | Grounded practical theory with interpretive analysis/ theme-oriented discourse analysis. |
| Demers (2013) | USA | Women (n=17)  Age range 22-43, median age 29 Branch: Army (n=10), Marines (n=3), Navy (n=4).  Rank: Enlisted (94%) | Van Gennep’s Rite of passage ^24^ | Focus groups with semi-structured interviews | Critical interpretive approach informed by hermeneutic phenomenology |
