## Supplementary material for "A Systematic Review of Military to Civilian Transition: The Role of Gender": Table 2 (from another version of this preprint: version 2, 2024-04-17)

| **Table 2: Qualitative Themes Identified in the Systematic Review** | | | | | | | | | | | | |
| --- | --- | --- | --- | --- | --- | --- | --- | --- | --- | --- | --- | --- |
|  | In Service Military Experience | | | | | | Post-Military Transition | | | | | |
| Articles | Military Identity | Military Norms | Feminine at odds with Masculine Ideal | Gender inequality:  Lack of equipment & services | Gender Discrimination & Misogyny | Military sexual harassment and sexual assault | | Loss of a military identity | Military Civilian Divide | Loss of Purpose | Female Veteran Identity | Civilian feminine norms at odds with military norms |
| Raabe et al. (2024) | X | X |  |  |  |  | | X | X | X |  |  |
| Barrington et al. (2023) | X | X | X |  | X |  | | X | X | X |  |  |
| Murray & Cancio (2023) | X | X |  |  |  |  | | X | X | X |  |  |
| Rattray et al. (2023) | X | X | X | X | X | X | | X | X | X | X | X |
| Barnett et al. (2022) | X | X |  |  |  |  | | X | X | X |  | X |
| Eichler (2022) | X | X | X | X | X | X | |  |  |  | X | X |
| Guthrie-Gowerm & Wilson-Menzfeld (2022) | X | X |  |  |  |  | | X | X |  |  | X |
| Laferty et al. (2022) |  | X |  | X | X | X | |  |  |  | X | X |
| Boros et al. (2021) | X | X | X |  | X | X | | X |  |  |  |  |
| Daphna-Tekoah et al. (2021) | X | X | X |  | X | X | |  |  |  |  |  |
| Sayer et al. (2021) | X | X |  |  | X | X | |  | X | X | X |  |
| Leigh & Koblinsky (2017) | X | X | X |  |  | X | |  |  |  |  | X |
| Libin et al. (2017) | X | X |  |  |  |  | | X |  |  |  |  |
| Orazem et al. (2017) | X | X |  |  |  |  | | X | X | X |  |  |
| Ahern et al. (2015) | X | X |  |  |  |  | | X | X | X |  |  |
| Burkhart & Hogan (2015) | X | X | X |  | X | X | | X | X | X |  | X |
| Mankowski et al. (2015) | X | X |  |  |  | X | |  |  |  |  |  |
| Koenig et al. (2014) | X | X |  |  |  |  | | X | X | X |  |  |
| Demers (2013) | X | X | X |  | X | X | | X | X |  |  | X |
