## Supplementary material for "A Systematic Review of Military to Civilian Transition: The Role of Gender": Figure 1 PRISMA (from another version of this preprint: version 2, 2024-04-17)

**Figure 1: PRISMA Flow Diagram**

**Identification of studies via databases and registers**

**Records removed *before* screening (n=15,614):**

Automated duplication removal (n=14,509)

Manual duplication removal

(n=1,105)

**Records identified from databases (n=25,727):**

Medline (n=2,492)

Embase (n=10,407)

PsychInfo (n=1,684)

Web of Science (n=10,407)

Pubmed (n=2,708)

EBSCO (n=3,300)

Global Health (n=95)

**Identification**

Records excluded

(n=9,952)

Records screened

(n=10,113)

Reports not retrieved

(n=0)

Reports sought for retrieval

(n=161)

**Screening**

Reports assessed for eligibility

(n=161)

**Reports excluded (n=142):**

Dissertation (n=18)

Not qualitative (n=109)

Insufficient data on women (n=8)

Transition from deployment (n=5)

National Guard only (n=2)

Studies included in review

(n=19)

**Included**

### *From:* Page MJ, McKenzie JE, Bossuyt PM, Boutron I, Hoffmann TC, Mulrow CD, et al. The PRISMA 2020 statement: an updated guideline for reporting systematic reviews. BMJ 2021;372:n71. doi: 10.1136/bmj.n71

For more information, visit: <http://www.prisma-statement.org/>
