## Supplementary material for "A Systematic Review of Military to Civilian Transition: The Role of Gender": JBI (from another version of this preprint: version 2, 2024-04-17)

| Joanna Briggs Institute (JBI) Qualitative Critical Appraisal Checklist for Qualitative Studies | | | | | | | | | | | |
| --- | --- | --- | --- | --- | --- | --- | --- | --- | --- | --- | --- |
|  | Q1 | Q2 | Q3 | Q4 | Q5 | Q6 | Q7 | Q8 | Q9 | Q10 | Q11 |
| *Raabe (2024)* | Yes | Yes | Yes | Yes | Yes | Yes | Yes | Yes | Yes | Yes | Yes |
| *Barrington et al. (2023)* | Yes | Yes | Yes | Yes | Yes | Yes | Yes | Yes | Yes | Yes | Yes |
| *Murray (2023)* | Yes | Yes | Yes | Yes | Yes | No | Yes | Yes | Yes | Yes | Yes |
| *Rattray (2023)* | Yes | Yes | Yes | Yes | Yes | Yes | Yes | Yes | Yes | Yes | Yes |
| *Barnett (2022)* | Yes | Yes | Yes | Yes | Yes | No | No | Yes | Yes | Yes | Yes |
| *Eichler (2022)* | Yes | Yes | Yes | Yes | Yes | No | No | Yes | Yes | Yes | Yes |
| *Guthrie-Gowerm (2022)* | Yes | Yes | Yes | Yes | Yes | No | Yes | Yes | Yes | Yes | Yes |
| *Laferty (2022)* | No | Yes | Yes | Yes | Yes | No | No | Yes | Yes | Yes | Yes |
| *Boros (2021)* | Yes | Yes | Yes | Yes | Yes | No | Yes | Yes | Yes | Yes | Yes |
| *Daphna-Tekoah (2021)* | Yes | Yes | Yes | Yes | Yes | Yes | Yes | Yes | Yes | Yes | Yes |
| *Sayer (2021)* | No | Yes | Yes | Yes | Yes | No | No | Yes | Yes | Yes | Yes |
| *Leigh (2017)* | Yes | Yes | Yes | Yes | Yes | No | No | Yes | Yes | Yes | Yes |
| *Libin (2017)* | Yes | Yes | No | No | Yes | No | No | No | Yes | Yes | Yes |
| *Orazem (2017)* | No | Yes | Yes | Yes | Yes | No | No | Yes | Yes | Yes | Yes |
| *Ahern (2015)* | Yes | Yes | Yes | Yes | Yes | No | No | Yes | Yes | Yes | Yes |
| *Burkhart (2015)* | Yes | Yes | Yes | Yes | Yes | No | No | Yes | Yes | Yes | Yes |
| *Mankowski (2015)* | No | Yes | Yes | Yes | Yes | No | No | Yes | Yes | Yes | Yes |
| *Koenig (2014)* | Yes | Yes | Yes | Yes | Yes | Yes | Yes | Yes | yes | Yes | Yes |
| *Demers (2013)* | Yes | Yes | Yes | Yes | Yes | No | No | Yes | Yes | Yes | Yes |

Q1: Is there congruity between the stated philosophical perspective and the research methodology?

Q2: Is there congruity between the research methodology and the research question or objectives?

Q3: Is there congruity between the research methodology and the methods used to collect data?

Q4: Is there congruity between the research methodology and the representation and analysis of data?

Q5: Is there congruity between the research methodology and the interpretation of results?

Q6: Is there a statement locating the researcher culturally or theoretically?

Q7: Is the influence of the researcher on the research, and vice- versa, addressed?

Q8: Are participants, and their voices, adequately represented?

Q9: Is the research ethical according to current criteria or, for recent studies, and is there evidence of ethical approval by an appropriate body?

Q10: Do the conclusions draw in the research report flow from the analysis, or interpretation, of the data?

Q11: Were the specific directives for new research appropriate?
