## Supplementary material for "A Systematic Review of Military to Civilian Transition: The Role of Gender": Themes_Subthemes (from another version of this preprint: version 2, 2024-04-17)

| **Appendix B: Themes and Subthemes** | | | |
| --- | --- | --- | --- |
| **Author / Publication Year** | **Country** | **Findings** | **Themes and subthemes** |
| Raabe et al.  (2024) | USA | Veterans face challenges in identifying relevant, purposeful employment opportunities, securing employment, and translating military skills to civilian contexts. Veterans had to adjust to differences in workplace norms between the civilian and military context. | 1. Veterans experienced different levels of relatedness in the military community during their transition 2. Perceptions of choice and control had varying influences on veterans’ sense of autonomy during their transition 3. Veterans experienced a false sense of competence starting the transition. 4. Figuring out how to transfer military experiences and skills to civilian jobs posed a meaningful challenge for veterans’ sense of competence throughout the application process 5. Multiple factors helped veterans regain their sense of competence during the job search 6. Veterans experienced difficulties adjusting to new civilian job environment 7. Family played a major role in fostering veterans’ sense of autonomy during the transition |
| Barrington et al. (2023) | UK | Veterans' military identity and experiences significantly impact their health and support-seeking behaviours upon transitioning to civilian life. Tailoring interventions to leverage veterans' unique military identities, especially through peer support, may improve support services for alcohol use. | 1. Searching for safety    1.1. Escaping unacceptance    1.2. Safey in structure 2. Healing with honour    2.1. Kin and kind    2.2. Healing through outrage    2.3. Compassionate flows |
| Murray & Cancio (2023) | USA | Reintegration into the civilian community for veterans who experienced burns during service was strengthened by peer support. Veterans also benefited from future oriented thinking and strong community (both civilian and veteran) network. | 1. Supportive community    1.1. Veteran specific support    1.2. Burn care    1.3. Peer relationships    1.4. Education-work-hobbies    1.5. Financial benefits 2. Future-oriented thinking    2.1. Turning point    2.2. Desire to serve    2.3. New meaning in life    2.4. Posttraumatic growth. |
| Rattray et al. (2023) | USA | Women veterans navigate complex and intersecting identities which are magnified by gender inequalities.  Early in the transition process, women veterans felt marginalized in accessing healthcare compared to their military spouses and male veteran peers. | 1. Reintegration 2. Social Network 3. Health 4. Career 5. Identity and self-perception |
| Barnett et al. (2022) | Australia | Programmes which offer supportive social networks and leverage veterans’ desire to give back to community may improve reintegration. Social mapping of veterans’ social group structures can help inform clinicians of veterans social groups. | 1. Military life, transition and identity    1.1. Forming military identities    1.2. Transition experiences and well-being 2. Identity, social groups and transitioning to civilian life 3. Military identity: values and serving community |
| Eichler (2022) | Canada | Gendered experiences for females are continuous from military service to the civilian environment. Military women adjust to male norm and masculine ideal of the military member during service yet their veteran status is often not recognized in the civilian environment. | 1. Gendered experience during the service    1.1. Having to conform to the masculine ideal and male norm    1.2. Facing gender discrimination and gender-based violence    1.3. Being a military spouse and mother while serving 2. Gendered post service    2.1. Being a woman Veteran: Not fitting the male and masculine Veteran norm    2.2. Standing out as a Veteran woman: Not fitting femininity norms    2.3. Women Veterans as spouses and mothers |
| Guthrie-Gowerm & Wilson-Menzfeld (2022) | UK | Transition out of the military resulted in feelings of social isolation and loneliness which hindered connections with civilians. These sentiments were found through the lifespan. | 1. A sense of loss    1.1. Formation of bonds    1.2. Loss of identity    1.3. Detached social networks 2. Difficulty connecting in civilian life    2.1. Different social norms    2.2. Experiential differences 3. Seeking out familiarity    3.1. Reconnecting to ex-military community    3.2. Connecting through shared interests |
| Laferty et al. (2022) | USA | Healthcare systems were inadequately prepared to address needs of female veterans. | 1. Military experience 2. Transitioning home    2.1. Emerging health concerns and development of chronic and complex conditions    2.2. Impacts on relationships    2.3. Challenges related to children and childcare    2.4. Employment and career difficulties 3. Seeking care 4. Coping and symptom management |
| Boros et al.  (2021) | USA | Gender influenced their in-service and transition experience. Women grappled with a post-military identity, grieving their military identify and composing a new identity. | 1. Family support    1.1. Before joining the military    1.2. After separating from the military 2. Mandatory conformity    2.1. Family    2.2. Military 3. Identity    3.1. Loss of identity    3.2. Gaining of identity 4. Service    4.1. Service after military service    4.2. Lessons learned 5. Gender inequality    5.1. Race intersection    5.2. Disability intersection    5.3. Pregnancy intersection 6. Symptoms    6.1. Physical    6.2. Emotional 7. Opportunities    7.1. Gained    7.2. Lost |
| Daphna-Tekoah et al. (2021) | USA / Israel | Veterans of both militaries expressed a need to be heard. Societies, therapists, and military institutions are not attuned to female veterans’ experiences and are not interested in what ails them. | 1. The battle to integrate and to prove themselves 2. Exposure to traumatic events and combat trauma 3. Sexual harassment and sexual assaults of combat women 4. Transition to civilian life |
| Sayer et al. (2021) | USA | Military-civilian divide and lack of public awareness isolate veterans. Individual characteristics in addition to social connections, community norms, interpersonal connection shape the reintegration experience. | 1. Military-civilian divide affects reintegration    1.1. Military culture and bonds    1.2. Veterans come back changed    1.3. Reintegration takes time 2. Bridging the military civilian divide    2.1. Recognize deployment hardships    2.2. Appreciate deployment accomplishments    2.3. Assist Veterans in getting help    2.4. Listen don’t judge    2.5. Recognize employment is critical |
| Leigh & Koblinsky (2017) | USA | Challenges in transitioning to a civilian environment included slower pace in civilian environment, adjusting to changes in children's development and managing difficult emotional interactions with family members. Women displayed resilience by making meaning of their military service, accessing veteran social support, and adapting on military acquired skills to the civilian environment. | Challenges   1. Incompatibility of military and civilian pace of life 2. Fear of intimacy 3. Missing children’s development 4. Family role changes 5. Family members as targets of anger 6. Sharing versus burdening family members 7. Family members’ lack of understanding the veteran’s emotional turmoil  Resilience strategies 1. Making meaning of military service 2. Restoring family rituals and routines 3. Accessing veteran support 4. Using military-acquired skills |
| Libin et al. (2017) | USA | Unplanned separation complicated reintegration. Veterans struggled with maintaining or fading one's military identity. | 1. Relationship with the Military: An untimely breakup 2. Relationship with civilian society: A world apart 3. Relationship with the VA: rehabilitation as a barrier and facilitator to re-entry |
| Orazem et al. (2017) | USA | Veterans struggled with Identity adjustment when reintegrating into a civilian environment. Veterans also reported difficulty finding meaning and purpose, missing the military lifestyle, and feeling left behind by civilians | 1. Feeling like one does not belong in civilian society 2. Missing the military culture and structured lifestyle 3. Holding negative views of civilian society 4. Feeling left behind compared to civilian counterparts 5. Having difficulty finding meaning in civilian world |
| Ahern et al. (2015) | USA (California) | Veterans report difficulty connecting with civilians, a lack of support from institutions, loss of identity and purpose on re-entering the civilian environment. | 1. Military as family    1.1. Caretaker    1.2. Structure 2. Normal is Alien    2.1. Disconnection    2.2. Unsupportive institutions    2.3. Lack of civilian structure    2.4. Loss of purpose 3. Searching for new normal    3.1. Support from a navigator    3.2. Embracing an ambassador role    3.3. Ease with time |
| Burkhart & Hogan (2015) | USA | Findings describe the process female veterans experience when transitioning from the military to civilian environment, examining the  stressors and adaptations of military life and gaining a dual identity of a veteran-civilian | 1. Choosing the military    1.1. Seeking opportunities    1.2. Pursuing adventure    1.3. Seeking safety. 2. 2. Adapting to the military    2.1. Experiencing cultural shock    2.2. Coping with cultural change    2.3. Becoming a member of the military 3. 3 Being in the military    3.1. Knowing military policies    3.2. Experiencing violence of war    3.3. Living for the moment    3.4. Acquiring value of camaraderie    3.5. Acquiring strong work ethic 4. Being a female in the military: Belonging to military health care    4.1. Being treated as an equal 5. Being a female in the military: Belonging to military outside of healthcare    5.1. Being treated as inferior/demeaned    5.2. Being a victim of sexual assault    5.3. Being betrayed    5.4. Becoming hardened 6. Experiencing stressors of being a civilian    6.1. Feeling unprepared for civilian life    6.2. Living two lives    6.3. Coping with PTSD 7. Making meaning of being a veteran-civilian    7.1. Belonging to a veteran-civilian community 1. Having pride in being a veteran-civilian |
| Mankowski et al. (2015) | USA | Women joined the military for opportunity and a desire to serve. The military was at times a hostile work environment and economic opportunities did not materialize for all women post service. | 2. Opportunity 3. Calling 4. Outcomes |
| Koenig et al. (2014) | USA | Veterans experience a "reverse culture shock" when transitioning to the civilian environment due to differences in norms and identities. Veterans identified three areas of tension, intrapersonal, professional/educational/ and interpersonal domains. Veterans offer strategies for managing these tensions. | 1. The intrapersonal domain 2. The professional/educational domain 3. Interpersonal domain 4. Cultural resources for resilience |
| Demers (2013) | USA (San Francisco, CA) | Conflict between military and civilian cultures and continual renegotiation between gender and identity | 1. Women at war    1.1. Becoming a soldier    1.2. Fighting two wars 2. Coming home    2.1. Dirty time bombs    2.2. Mourning who I was    2.3. Questioning who I am    2.4. Composing who I will be |
