## Supplementary material for "A Systematic Review of Military to Civilian Transition: The Role of Gender": Search Strategy (from another version of this preprint: version 2, 2024-04-17)

| **Ovid Medline** | | | | |
| --- | --- | --- | --- | --- |
| **Category** | **Search Num** | **Ovid Medline** | **Number (Feb 2023)** | **Number (Feb 2024)** |
| ex-Service members | 1 | exp Veteran/ | 21,310 | 22,793 |
|  | 2 | veteran*.mp | 43,184 | 46,184 |
|  | 3 | ex-service*.mp. | 133 | 142 |
|  | 4 | ex-force*.mp. | 5 | 5 |
|  | 5 | ex-military*.mp. | 36 | 40 |
|  | 6 | ex-soldier*.mp. | 17 | 19 |
|  | 7 | ex-Armed Force*.mp. | 8 | 8 |
|  | 8 | military veteran*.mp. | 1,918 | 2,177 |
|  | 9 | retired military personnel*.mp. | 33 | 36 |
|  | 10 | 1 - 9 (OR) | 46,234 | 49,289 |
| Women | 11 | exp Women/ | 43,407 | 45,553 |
|  | 12 | woman*.mp. | 257,556 | 269,645 |
|  | 13 | women*.mp. | 1,183,176 | 1,247,574 |
|  | 14 | female/ | 9,543,971 | 9,710,032 |
|  | 15 | female*.mp | 9,774,000 | 9,966,996 |
|  | 16 | service-woman*.mp. | 2 | 3 |
|  | 17 | service-women*.mp. | 147 | 158 |
|  | 18 | exp Gender Identity/ | 23,385 | 27,875 |
|  | 19 | gender*.mp. | 431,525 | 464,400 |
|  | 20 | feminin*.mp. | 7,907 | 8,370 |
|  | 21 | 11 - 20 (OR) | 10,046,833 | 10,272,898 |
| Transition general | 22 | Transition*.mp. | 533,758 | 572,884 |
|  | 23 | Adjust*.mp. | 260,384 | 275,963 |
|  | 24 | readjust*.mp | 3,354 | 3,501 |
|  | 25 | Reintegrat*.mp. | 4,712 | 5,060 |
|  | 26 | integrat*.mp | 694,314 | 767,100 |
|  | 27 | Resettle*.mp. | 2,689 | 2,917 |
|  | 28 | settle*.mp | 30,292 | 32,551 |
|  | 29 | release*.mp. | 818,368 | 868,067 |
|  | 30 | post military*.mp. | 46 | 59 |
|  | 31 | post service*.mp. | 109 | 123 |
|  | 32 | after service*.mp. | 229 | 248 |
|  | 33 | transition to civilian.mp | 66 | 73 |
|  | 34 | adapt*.mp. | 844,894 | 917,545 |
|  | 35 | 22-35(OR) | 2,394,062 | 2,542,273 |
| Ex-service members  +  Women  +  Transition | 36 | 10 and 21 and 35 ( Service Members + Women + Transition) | 2,474 | 2,492 |

| **Ovid Embase** | | | | |
| --- | --- | --- | --- | --- |
| **Category** | **Search Num** | **Ovid Embase** | **Number (Feb 2023)** | **Number (Feb 2024)** |
| ex-Service members | 1 | exp Veteran/ | 34,916 | 36,379 |
|  | 2 | veteran*.mp | 59,941 | 62,004 |
|  | 3 | ex-service*.mp. | 192 | 134 |
|  | 4 | ex-force*.mp. | 5 | 5 |
|  | 5 | ex-military*.mp. | 49 | 46 |
|  | 6 | ex-soldier*.mp. | 31 | 17 |
|  | 7 | ex-Armed Force*.mp. | 6 | 6 |
|  | 8 | military veteran*.mp. | 2,350 | 2,600 |
|  | 9 | retired military personnel*.mp. | 52 | 46 |
|  | 10 | 1 - 9 (OR) | 62,855 | 64,704 |
| Women | 11 | exp female/ | 11,615,610 | 11,897,134 |
|  | 12 | woman*.mp. | 476,523 | 477,619 |
|  | 13 | women*.mp. | 1,659,044 | 1,718,176 |
|  | 14 | female/ | 11,529,723 | 11,841,523 |
|  | 15 | female*.mp | 11,827,869 | 12,138,469 |
|  | 16 | service-woman*.mp. | 3 | 5 |
|  | 17 | service-women*.mp. | 181 | 190 |
|  | 18 | exp Gender Identity/ | 20968 | 23,329 |
|  | 19 | exp Gender | 344,050 | 367,818 |
|  | 20 | gender*.mp. | 721,937 | 786,645 |
|  | 21 | feminin*.mp. | 11,244 | 10,909 |
|  | 22 | exp femininity/ | 2,917 | 2,924 |
|  | 22 | 11 - 22 (OR) | 12,236,711 | 12,531,604 |
| Transition general | 23 | Transition*.mp. | 675,488 | 730,617 |
|  | 24 | Adjustment*.mp. | 340,022 | 360,902 |
|  | 25 | exp adjustment/ | 4,932 | 4,978 |
|  | 26 | readjust*.mp | 4,856 | 4,567 |
|  | 27 | Reintegrat*.mp. | 7,213 | 7,363 |
|  | 28 | integrate*mp | 492,848 | 545,277 |
|  | 29 | Resettle*.mp. | 3,124 | 3,084 |
|  | 30 | release*.mp. | 1,272,400 | 1,306,659 |
|  | 31 | post military*.mp. | 52 | 60 |
|  | 32 | post service*.mp. | 160 | 180 |
|  | 33 | after service*.mp. | 298 | 315 |
|  | 34 | transition to civilian.mp | 67 | 73 |
|  | 35 | adapt*.mp. | 977,340 | 1,025,933 |
|  | 36 | 24-36 (OR) | 3,582,227 | 3,775,147 |
| Ex-service members  +  Women  +  Transition | 37 | 10 and 22 and 36 ( Service Members + Women + Transition) | 4,599 | 5,035 |

| **PsycInfo** | | | | |
| --- | --- | --- | --- | --- |
| **Category** | **Search Num** | **PsycInfo** | **Number**  **(Feb 2023)** | **Number**  **(Feb 2024)** |
| ex- Service members | 1 | exp Veteran/ | 15,855 | 17,188 |
|  | 2 | veteran*.mp | 24,926 | 26,487 |
|  | 3 | ex-service*.mp. | 111 | 116 |
|  | 4 | ex-force*.mp. | 5 | 5 |
|  | 5 | ex-military*.mp. | 38 | 44 |
|  | 6 | ex-soldier*.mp. | 37 | 37 |
|  | 7 | ex-Armed Force*.mp. | 5 | 5 |
|  | 8 | military veteran*.mp. | 1,913 | 2,175 |
|  | 9 | retired military personnel*.mp. | 21 | 22 |
|  | 10 | 1 - 9 (OR) | 26,537 | 28,140 |
| Women | 11 | human females/ | 155,891 | 162,661 |
|  | 12 | woman*.mp. | 46,150 | 47,731 |
|  | 13 | women*.mp. | 322,392 | 339,801 |
|  | 14 | female*.mp | 1,149,692 | 1,184,642 |
|  | 15 | service-woman*.mp. | 5 | 5 |
|  | 16 | service-women*.mp. | 63 | 66 |
|  | 17 | exp Gender Identity/ | 44421 | 49,593 |
|  | 18 | gender*.mp. | 266,027 | 283,850 |
|  | 19 | feminin*.mp. | 17,674 | 18,391 |
|  | 20 | exp femininity/ | 5,250 | 5,507 |
|  | 21 | 11 - 20(OR) | 1,433,242 | 1,488,041 |
| Transition general | 22 | Transition*.mp. | 89,234 | 95,566 |
|  | 23 | Adjustment*.mp. | 136,106 | 140,946 |
|  | 24 | exp adjustment/ | 56,125 | 58,166 |
|  | 25 | readjust*.mp | 4,628 | 4,755 |
|  | 26 | Reintegrat*.mp. | 5,739 | 6,195 |
|  | 27 | integrate*mp | 130,447 | 139,135 |
|  | 28 | Resettle*.mp. | 2,288 | 2,508 |
|  | 29 | release*.mp. | 54,888 | 56,825 |
|  | 30 | post military*.mp. | 73 | 85 |
|  | 31 | post service*.mp. | 160 | 121 |
|  | 32 | after service*.mp. | 156 | 173 |
|  | 33 | transition to civilian.mp | 133 | 154 |
|  | 34 | adapt*.mp. | 265,434 | 282,055 |
|  | 35 | 22-34 (OR) | 630,371 | 665,917 |
| Ex-service members  +  Women  +  Transition | 36 | 10 and 21 and 35( Service Members + Women + Transition) | 1,594 | 1,684 |

| **Global Health** | | | | |
| --- | --- | --- | --- | --- |
| **Category** | **Search Num** | **Global Health** | **Number**  **(Feb 2023)** | **Number**  **(Feb 2024)** |
| ex-service members | 1 | veteran*.mp | 5,566 | 6,001 |
|  | 2 | ex-service*.mp. | 22 | 23 |
|  | 3 | ex-force*.mp. | 0 | 0 |
|  | 4 | ex-military*.mp. | 3 | 5 |
|  | 5 | ex-soldier*.mp. | 8 | 8 |
|  | 6 | ex-Armed Force*.mp. | 0 | 0 |
|  | 7 | military veteran*.mp. | 275 | 300 |
|  | 8 | retired military personnel*.mp. | 8 | 8 |
|  | 9 | 1 - 9 (OR) | 5,600 | 6,038 |
| Women | 10 | female*.mp | 246,851 | 267,007 |
|  | 11 | woman*.mp. | 30,788 | 32,113 |
|  | 12 | women*.mp. | 379,309 | 412,626 |
|  | 13 | service-woman*.mp. | 2 | 2 |
|  | 14 | service-women*.mp. | 52 | 59 |
|  | 15 | gender Identity.mp | 683 | 906 |
|  | 16 | gender*.mp. | 95,964 | 105,126 |
|  | 17 | feminin*.mp. | 786 | 845 |
|  | 18 | 11 - 20(OR) | 622,730 | 673,031 |
| Transition general | 19 | Transition*.mp. | 34134 | 37,661 |
|  | 20 | Adjustment*.mp. | 51350 | 54,790 |
|  | 21 | readjust*.mp | 289 | 313 |
|  | 22 | Reintegrat*.mp. | 488 | 544 |
|  | 23 | integrate*mp | 60,070 | 67,641 |
|  | 24 | Resettle*.mp. | 1,087 | 1,164 |
|  | 25 | release*.mp. | 85,643 | 92,743 |
|  | 26 | post military*.mp. | 10 | 11 |
|  | 27 | post service*.mp. | 21 | 23 |
|  | 28 | after service*.mp. | 53 | 57 |
|  | 29 | transition to civilian.mp | 11 | 11 |
|  | 30 | adapt*.mp. | 83,803 | 92,968 |
|  | 31 | 22-34 (OR) | 304,842 | 334,264 |
| Ex-service members  +  Women  +  Transition | 33 | 10 and 18 and 31( Service Members + Women + Transition) | 95 | 101 |

| **Web of Science** | | | |
| --- | --- | --- | --- |
| **Category** | **Web of Science:** | **Number**  **(Feb 2023)** | **Number**  **(Feb 2024)** |
| ex-service members | AK=(veteran* ) OR TI=(veteran*) OR AB= (veteran*)  OR AK=(ex-service*) OR TI =(ex-service*) or AB=(ex-service*)  OR AK=(ex-military*) OR TI=(ex-military*) OR AB= ( ex-military*)  OR AK=(ex-soldier*) OR TI=(ex-soldier*) OR AB= (ex-soldier*)  OR AK=(ex-armed force*) OR TI=(ex-armed force*) OR AB= ( ex-armed force* )  OR AK=(military veteran*) OR TI=(military veteran*) OR AB= (military veteran*)  OR AK=(retired military person*) OR TI=(military person* ) OR AB= ( military person*) | 67,361 | 116,023 |
| Women | ALL=(female*) OR ALL=(woman*) OR ALL=(women*) OR ALL=(service-women*) OR ALL=(service-woman*) OR ALL=(gender identity*) OR ALL=(gender*) OR ALL=(feminin*) | 3,709,224 | 13,813,317 |
| Transition general | ALL=(Transition*) OR ALL=(Adjustment*) OR ALL=(readjust*) OR ALL=(Reintegrat*) OR ALL=(integrate*) OR ALL=(Resettle*) OR ALL=(release*) OR ALL=(post military*) OR ALL=(post service*) OR ALL=(after service*) OR ALL=(transition to civilian) OR ALL=(adapt*) | 7,558,092 | 11,647,697 |
| Ex-service members  +  Women  +  Transition | ( Service Members + Women + Transition) | 2,914 | 10,407 |

| Pubmed February 2023 | | |  |
| --- | --- | --- | --- |
| Category | Terms | Numbers (February 2023) | Numbers (February 2024) |
| Ex-service members | "veteran*"[TW] OR "ex service*"[TW] OR "ex force*"[TW] OR "ex military*"[TW] OR "ex soldier*"[TW] OR "ex armed force*"[TW] OR "military veteran*"[TW] OR "retired military personnel*"[TW] | 50,161 | 53,628 |
| Women | "woman*"[TW] OR "women*"[TW] OR "female*"[TW] OR "service woman*"[TW] OR  "service women*"[TW] OR "gender identity*"[TW] OR "gender*"[TW] OR "feminin*"[TW] | 10,019,990 | 10,269,743 |
| Transition | "transition*"[TW] OR "Adjust"[TW] OR "readjust"[TW] OR "reintegrat*"[TW] OR "integrat*"[TW] OR "resettle*"[TW] OR "settle*"[TW] OR "release*"[TW] OR "post military*"[TW] OR "post service*"[TW] OR "after service*"[TW] OR "transition to civilian"[TW] OR  "adapt*"[TW] | 2,817,815 | 3,044,631 |
| Ex-service members  +  Women  +  Transition | (ex-Service Members + Women + Transition) - TW | 2,477 | 2,708 |

| EBSCO: February 2023, update February 2024 | | |  |
| --- | --- | --- | --- |
|  | terms | Numbers (February 2023) | Numbers (February 2024) |
| Ex-service members | TI Veteran* OR AB veteran* OR TI ex-service OR AB ex-service OR TI ex-military* OR AB ex-military* OR TI ex-soldier* OR AB ex-soldier* OR TI ex-Armed Force* OR AB ex-Armed Force* OR TI military veteran* OR AB military veteran* OR TI retired military personnel* OR AB retired military personnel | 27,823 | 30,532 |
| Women | Search: (woman*) OR (women*) OR (female*) OR (service-woman*) OR (service-women*) OR (Gender Identity*) OR (gender*) OR (feminin*) | 2,458,318 | 2,644,789 |
| Transition | Search: (Transition*) OR (Adjust*) OR (readjust*) OR (Reintegrat*) OR (integrat*) OR (Resettle*) OR (settle*) OR (release*) OR (post military*) OR (post service*) OR (after service*) OR (transition to civilian) OR (adapt*) | 764,194 | 929,402 |
| Ex-service members  +  Women  +  Transition | ( Service Members + Women + Transition) | 3,199 | 3,300 |
