## Supplementary material for "A Systematic Review of Military to Civilian Transition: The Role of Gender": Quotes (from another version of this preprint: version 2, 2024-04-17)

| **Appendix D: Selected Quotes** | | |
| --- | --- | --- |
| Themes | | Quotes |
| In Service Military Experience | Military Identity | “I had to [enlist], duty and self-sacrifice and all that, I don’t know how to describe it, I couldn’t explain it, I’d have done it for free.” (Mankowski et al., 2015) pg.319 |
|  | Military Norms | “You eat a certain way, you talk a certain way, you walk a certain way.” (officer) (Burkhart & Hogan, 2015)pg . 115  “being yelled at,” “harassed,” “Culture shock ... very mentally tough ... there’s always someone telling you you’re horrible, you’re a magot” (enlisted) (Burkhart & Hogan, 2015)pg.115 |
|  | Feminine at odds with Masculine Ideal | “In the military, guys can be crude, and I’d tell them [male soldiers] - say whatever you want just like if I was a male-, and I would try to fit in [by] being like a guy as much as I could just for that reason; because if you can play well, you can work well. (Demers, 2013) pg. 499  “. . . It was nonstop . . . They didn’t want me to succeed . the fact that I could do what they were doing, and do it well, took away from their masculinity or the manliness of what they were doing” (Demers, 2013)pg. 500 |
|  | Lack of equipment & services | “They just weren’t prepared for the amount of women, I think in the military at that point.” (Lafferty et al., 2022) pg. 3  “Yeah, the rucksack that we had to wear and that sort of thing ’cause it never ever fit masculine ideal and male norm right. And that’s, my back problem is because of that.” (Eichler, 2022) pg. 40 |
|  | Gender Discrimination & Misogyny | “In the military they would say that in order for you to be a strong man you can’t be a weak woman, so they’re building strength up from saying that, you know, strength is the opposite of anything that is feminine. So, there was a lot of stuff that was happening along those lines that was a difficult pill to be able to swallow, but so, just having to deal with that level of violence directed toward me as a female was a hard thing to have to deal with.” (Eichler, 2022) pg. 40  The females have to try harder to be better and stronger and faster than the males just to be considered even equal. (Daphna-Tekoah et al., 2021) pg. 6 |
|  | Military sexual harassment and sexual assault | “I mean, Iraq—you expect people to shoot at you, you expect people to die, you expect people to be killed. You don't expect your fellow soldiers to turn on you.” (Ahern et al., 2015) pg. 5  “I mean, oh my God, the harassment just doesn’t stop, and it’s just the environment of the way men think or they’re… encouraged to think because they’re military.” (Lafferty et al., 2022) pg. 3 |
| Post-Military Transition | Loss of a military identity | “When I first left, I was totally and utterly bereft. The day I handed my ID card in I sat and wept because that was my whole identity. . .” (Guthrie-Gower & Wilson-Menzfeld, 2022) pg. 7  “My entire identity was caught up in my military career, Being a civilian is so unknown to me and I still struggle to really feel like I belong in that ‘club’” (Orazem et al., 2017) pg. 7  “the military is not just a profession, it’s a way of life… the way that you act, even the morals that you’re bound by are from a certain code.”… “you can’t fully transition… You might physically be there, but your heart doesn’t transition.” (Raabe et al., 2024) pg. 52 |
|  | Military Civilian Divide | “. . .I almost felt that when I first got out, I couldn’t speak freely I had to watch everything that I said. So that kind of silences you a little bit.” (Guthrie-Gower & Wilson-Menzfeld, 2022) pg. 9  “I was never alone, I was never isolated without anyone, but I could feel lonely in a crowded room. I could have felt absolutely lonely because at the time there was no one that got me, nobody who understood me and therefore nobody I could talk to about what, you know, things that were playing on my mind. . .” (Guthrie-Gower & Wilson-Menzfeld, 2022) pg. 10  “Reintegration to civilian life is an exceptional challenge. The veteran has made an investment to one job and is now beginning an entirely new sector. the civilian sector has new rules, new requirements, and even and a new ‘uniform’”. (Sayer et al., 2021) pg. 401  “You don’t find a whole lot of queen bees in the military like you do in the civilian sector—by queen bee I’m just talking about that woman that makes it to the top and then [is] not willing to help other women.” (Rattray et al., 2023) pg. 5 |
|  | Loss of Purpose | “It’s really hard to put in words but I just miss the environment. I miss the common goals... the way people put aside their own personal [agendas]... I used to run a lot and [the] feeling is just like that...‘yeah man, let’s go do it!’ I’ve never really gotten the same thing on the civilian side, even though I try and pour my heart into things” (Ahern et al., 2015) pg. 6  “I miss the sense of purpose. I miss the sense of worth. Yes, everyone became and left different than what we were when we arrived. But yes, we made a difference. There is no resuming civilian life after that.” (Orazem et al., 2017) pg. 8  “It’s giving back [to community]. I think that’s been the biggest reward. I’ve had first-class indoctrination as to what serving the community means. That’s the reward, I think, finding ways to continue that, just not in uniform” (Barnett et al., 2022) pg. 1031 |
|  | Female Veteran Identity | “As a female in the military, ex-military, and the VA world, women need to be advocates for women. Because there’s all these services but most all of them are all geared around men, not that they are gender labelled, but it’s just known that most men, that men are in the military.” (Lafferty et al., 2022) pg. 5 |
|  | Civilian feminine norms at odds with military norms | “Well first it would be like people not understanding why a woman in the first place would want to go in the military so there’s an assumption that there must be something wrong with me, right? So, there’s just that part. And then the, again, the Rambo factor, so, oh, you know, she would have been in command and control, and she would have, so she reacted that way and that’s why she’s psycho, she’s so aggressive, ex-military. And then you go like, no, everybody sort of reacts that way and then it was like, yeah, being like, you know the, what they would expect with the whole feminine thing, and I don’t think I’m not feminine, I just think that I was a little bit more kind of assertive in how sort of I handled myself.” (Eichler, 2022) pg. 40 |
